# Diagnostic Accuracy of a locally deployed Large Language Model Algorithm for Automated Code Stroke Pathway Identification in Emergency Department Triage Notes

**DOI:** 10.64898/2026.08.03.26359639

**Authors:** Michael Valente, Angelos Sharobeam, Wing Kiu Chan, Jason Vuong

## Abstract

**Background:** Delayed Code Stroke activation contributes to worse outcomes in acute stroke. Emergency Department (ED) triage notes contain free-text clinical information that could enable automated, real-time pathway activation. We evaluated the diagnostic accuracy of a multi-pass large language model (LLM) pipeline for identifying patients meeting Code Stroke criteria from ED triage notes.

**Methods:** A retrospective cross-sectional study was conducted at Monash Medical Centre, Melbourne, Australia. De-identified triage notes from 3,023 ED presentations over a one-month period (September–October 2023) were analysed. The pipeline applied sequential passes for translation, stroke symptom identification, mimic exclusion, baseline functional status, temporal window classification, and symptom resolution. Six locally deployed language models were evaluated. Performance was assessed against two reference standards: neurologist-labelled diagnosis and documented ED Code Stroke activation. Primary outcomes were sensitivity and specificity; secondary outcomes included PPV, NPV, and Gwet’s AC1. Reliability of the neurologist reference standard was assessed by blinded independent re-review of a stratified random sample of 200 presentations by a second neurologist.

**Results:** Of 3,023 presentations, 136 were neurologist-labelled positive. Agreement between the primary and a blinded second neurologist on a 200-note reliability sub-sample was almost perfect (raw agreement 95.0%, Cohen’s κ 0.900, 95% CI 0.838–0.959). The cohort included 140 ED Code Stroke activations (median age 69, IQR 56–81 years), of whom 83 (59.2%) had confirmed stroke diagnosis. Sixteen patients (11.4%) underwent endovascular clot retrieval and 4 (2.9%) received thrombolysis. The best-performing model (Qwen 2.5 14B) achieved sensitivity 0.890 (95% CI 0.826–0.932), specificity 0.993 (0.989–0.996), PPV 0.858 (0.791–0.906), and NPV 0.995 (0.991–0.997). Pairwise McNemar testing demonstrated statistically superior overall accuracy for Qwen 2.5 14B over Llama 3.1 8B, Phi-4 14B, and Mistral 3 14B (all p<0.001 after Holm correction), with no significant difference detected versus Nemotron-Nano-12B-v2 or Qwen 3 14B.

**Conclusions:** A locally deployed language model demonstrates acceptable sensitivity and specificity for automated Code Stroke identification from free-text triage notes. Performance was comparable across the two best models, suggesting that capable open-weight models in this parameter range may be sufficient to proceed with ongoing internal testing and external validation. The pipeline operates without internet connectivity or model retraining on patient data, supporting feasibility for real-world ED integration.

## Introduction

Stroke is a leading cause of disability worldwide, and early identification is strongly linked to improved functional outcome[1]. Despite the implementation of systems of care such as the F.A.S.T. algorithm[2], delays to treatment persist at multiple stages in the identification and treatment chain. The potential for improvement at each stage remains significant: every hour of delay to reperfusion of a vessel occlusion can result in a 5% lower likelihood of returning to independent function[3]. Similarly, early bundled treatment with blood pressure lowering in haemorrhagic stroke has been shown to improve outcome[4].

Emergency triage represents a critical step in this chain. Even with appropriate training, approximately 10-20% of stroke cases are estimated to be missed at the emergency department triage step[5, 6]. Patients presenting with aphasia, posterior circulation symptoms, or those from culturally and linguistically diverse backgrounds are disproportionately affected[5, 6]. Triage also serves as the key handover point between paramedical assessment and in-hospital stroke code activation. Failures at this step, whether missed diagnoses or downstream treatment delays, have been identified as a persistent limitation of traditional systems.

Large language models present a promising approach to assist with early and automated stroke identification at triage. By processing unstructured clinical text, these models can identify the presence of stroke symptoms and extract clinically relevant information such as time of onset from paramedic handover notes. Deploying a lightweight, on-device model is particularly advantageous in this setting. Unlike cloud-based or fine-tuned alternatives, on-device inference avoids transmitting patient data over the internet and does not require training processes that risk internalising private health information within the model[7].

However, as with any screening tool, sufficient sensitivity and specificity must be demonstrated to ensure clinical utility. Hallucination (where model output diverges from the input data) is a recognised failure mode of large language models, particularly at smaller parameter sizes[8]. Strategies such as prompt engineering[9], structured prompt chaining[10], and constrained decoding[11] have been explored to mitigate this risk. This study evaluates the performance of a lightweight, on-device large language model for automated stroke identification from emergency triage data, assessing its diagnostic accuracy and examining the effect of inference strategies on reliability.

## Methods

### Study Design

A retrospective cross-sectional diagnostic accuracy study was conducted at Monash Medical Centre, Melbourne, Australia. The study was approved by the Monash Health Human Research Ethics Committee A (QA/127358/MonH-2026-524757). De-identified triage notes from 3,023 consecutive emergency department presentations over a one-month period (September–October 2023) were included. Triage notes for patients younger than the age of 18 were excluded. All other triage notes recorded during the study period were analysed regardless of presenting complaint to capture the full case-mix this system would encounter after deployment.

### Pipeline Overview

The automated classification pipeline was developed to determine whether a given ED triage note met criteria for Code Stroke activation. Code Stroke criteria were implemented as historically accurate for the study period at Monash Medical Centre. This included: CAT_1 (highest-priority activation, corresponding to presentation within 9 hours or unknown onset), CAT_2 (presentation at 9–24 hours with ongoing symptoms), or NO_CODE (activation not warranted). Patients with advanced dementia or presenting from high-level care nursing facilities were excluded as per the stroke criteria. The first 100 cases were initially withheld to perform rapid iteration and benchmarking of engineering steps. These cases were not included in final analysis.

The resulting pipeline processed each note through a series of sequential clinical reasoning steps, implemented as independent calls to a locally deployed large language model (Figure 1). Each step addressed a discrete clinical question: whether acute focal neurological symptoms were described, whether findings were better explained by a recognised stroke mimic, whether baseline functional exclusions applied, whether the symptom onset fell within a treatment-eligible time window, and whether symptoms had fully resolved. The outputs of each step were combined by deterministic classification logic (implemented in Python) to assign a final category: CAT_1, CAT_2 or NO_CODE.

**Figure 1:**
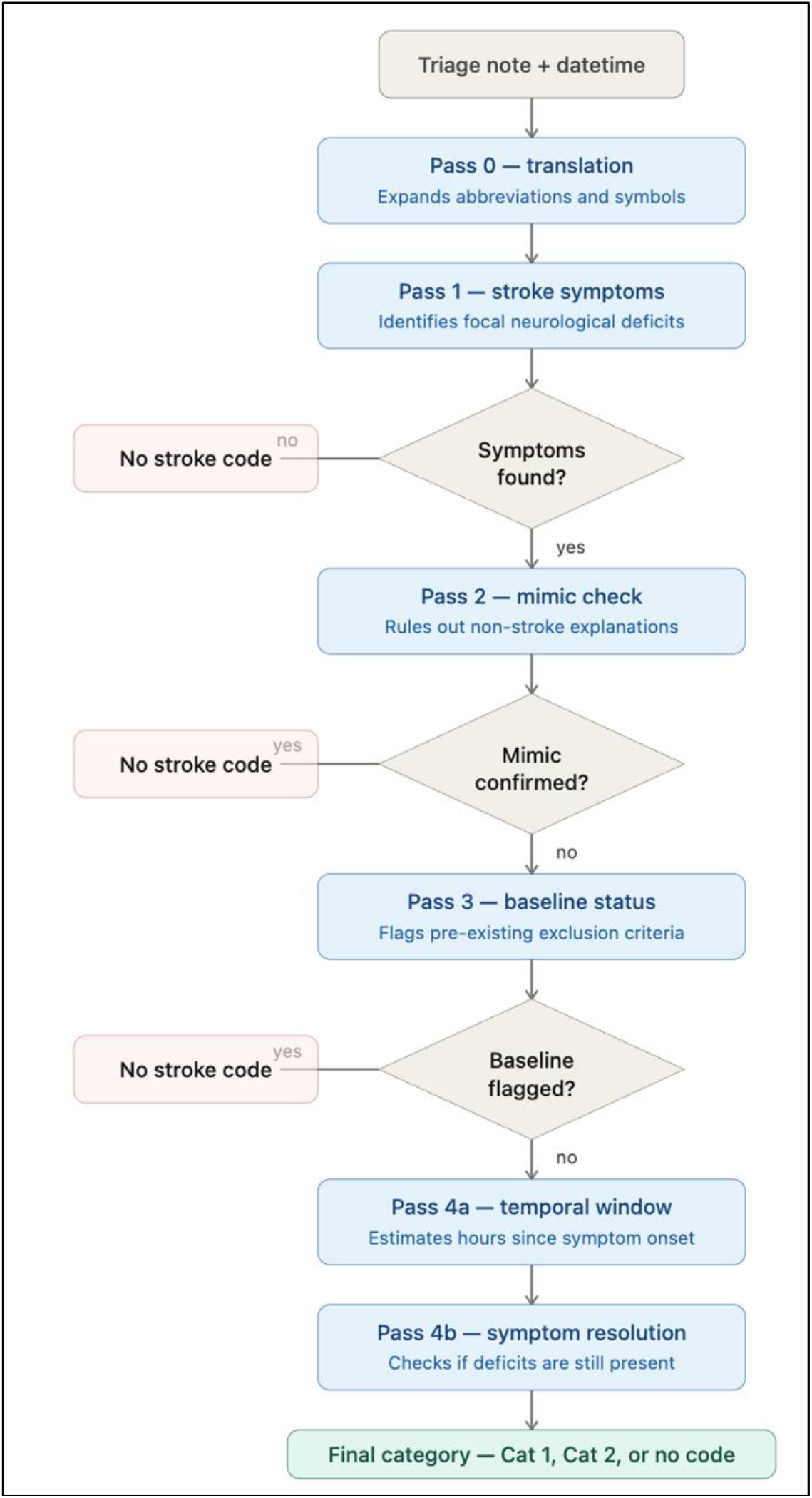
Stroke identification algorithm prompt-chain summary.

The pipeline was designed for fully local deployment with no internet connectivity required during operation. All model inference occurred on-device via the Ollama inference server, with no patient data transmitted to external servers. The use of pre-trained general-purpose models (as opposed to models fine-tuned on clinical data) ensured that no patient information was incorporated into model weights during any training process. This architecture addresses key privacy and data sovereignty requirements for clinical deployment.

### Reference Standards

Pipeline performance was assessed against two reference standards. The primary reference standard was neurologist determination of Code Stroke eligibility. A stroke neurologist (MV, 10 years’ experience) retrospectively reviewed each de-identified triage note, blinded to documented ED Code Stroke activation status, final clinical diagnoses, and all pipeline outputs, and assigned a binary label (Code Stroke–eligible or not) according to the same written institutional criteria used by the pipeline (including symptom presence, mimic exclusion, baseline functional status, time window, and symptom resolution). The secondary reference standard was documented ED Code Stroke activation, representing the existing clinical pathway decision as recorded in real time by the treating emergency team.

Reliability of the primary reference standard was assessed in a blinded inter-rater sub-study. A second board-certified neurologist (with 10 years’ stroke experience), blinded to the original labels, to the pipeline outputs and to documented ED Code Stroke activation status, independently re-reviewed a sample of presentations and assigned the same binary stroke label using the same written labelling rules.

The sub-study sample size was determined a priori using the method described by Bujang and Baharum for kappa-based reliability studies[12]. Assuming an expected Cohen’s kappa of 0.80, balanced class prevalence of 0.50, a two-sided alpha of 0.05, and a target 95% confidence interval half-width of ±0.10, the minimum required sample was 96 cases; 200 notes were sampled to provide additional precision.

Because the prevalence of neurologist-labelled stroke in the full cohort was 4.5%, simple random sampling would have yielded too few positive cases for stable estimation of agreement. Stratified random sampling was therefore used: 100 notes were drawn at random from those originally labelled stroke-positive and 100 from those originally labelled stroke-negative.

### Prompt Engineering Strategies

The classification task was decomposed into discrete sub-tasks (prompt chaining), each addressed by an independent model call with an empty conversational context. For steps requiring nuanced interpretation (symptom identification, baseline assessment, and resolution assessment), chain-of-thought prompting was used: the model first produced step-by-step reasoning in free text, which was then supplied as context to a second call that returned a structured JSON classification.

All steps used greedy decoding (temperature 0) to ensure reproducibility, apart from the translation step (temperature 0.1). As a preprocessing step, each triage note was first rewritten by the model into plain English, expanding clinical abbreviations and Australian shorthand (e.g. “2/7” meaning two days); all subsequent steps operated on the translated text. Model outputs were parsed deterministically, with parse failures defaulting conservatively toward non-activation. Early-exit logic terminated processing when a note could be conclusively classified without completing all steps.

### Models Evaluated

Six open-weight language models were evaluated in the primary comparison, all deployed locally via the Ollama inference server deployed on a single consumer-grade GPU: Llama 3.1 8B (Meta), Mistral 3 14B (Mistral AI), Nemotron-Nano-12B-v2 (NVIDIA), Phi-4 14B (Microsoft), Qwen 2.5 14B (Alibaba), and Qwen 3 14B (Alibaba). Models ranged from 8 to 14 billion parameters. All models were served using 4-bit quantisation. All models were used as pre-trained general-purpose models without fine-tuning or domain-specific training.

### Secondary Comparison: Prompt Chaining versus Single-Pass Inference

A secondary analysis compared three inference strategies to quantify the contribution of the prompt chaining architecture to diagnostic performance. This comparison was conducted across three models (Llama 3.1 8B, Qwen 2.5 14B, and Qwen 3 14B):

#### Multi-pass chain (primary configuration)

The full sequential pipeline as described above, with each clinical reasoning step addressed by a dedicated model call at temperature 0 with thinking mode disabled.

#### Single-pass inference

All clinical criteria (translation, symptom identification, mimic exclusion, baseline assessment, temporal analysis, resolution assessment, and final categorisation) were presented in a single consolidated prompt. The model was asked to reason through each step sequentially and produce a structured output in one inference call. Temperature and thinking mode settings matched the chain configuration.

#### Single-pass with thinking mode (Qwen 3 14B only)

The single-pass configuration was repeated with the model’s internal reasoning mode enabled and non-greedy sampling (temperature 0.6, top-p 0.95, following the model developer’s guidance). In this configuration the model produced an internal reasoning trace before its visible answer. Run-to-run determinism was lost due to the non-zero temperature.

### Statistical Analysis

For each model and reference standard combination, a two-by-two contingency table was constructed from the binary model prediction (CAT_1 or CAT_2 versus NO_CODE) and the binary reference label. Sensitivity, specificity, positive predictive value, and negative predictive value were calculated with Wilson score 95% confidence intervals. Balanced accuracy was computed as the arithmetic mean of sensitivity and specificity with a normal-approximation 95% confidence interval. Agreement between each model’s predictions and the reference standard was quantified using Cohen’s kappa and Gwet’s AC1. AC1 was included because kappa is known to produce paradoxically low values under high agreement with prevalence imbalance, which was expected given the low base rate of stroke presentations in an undifferentiated ED cohort.

To compare diagnostic performance across models, each patient was scored as correctly or incorrectly classified by each model against the reference standard. For every pair of models, a discordance table was constructed counting the patients where one model was correct and the other incorrect. McNemar’s test was applied to each pair to assess whether the discordance was symmetric (i.e. whether one model was systematically more accurate than the other). The chi-squared approximation was used for the full-sample comparison. Holm correction was applied across all model pairs within each analysis to control the family-wise error rate.

Agreement between the two neurologists on the reliability sub-sample was quantified as Cohen’s kappa with bootstrap 95% confidence intervals based on 2,000 resamples. McNemar’s exact test was used to assess whether disagreement between the two raters was systematically directional. Agreement was assessed on the composite binary stroke label; component-level agreement for the individual pipeline criteria was not assessed.

Missing model predictions were imputed as model-negative, representing the conservative direction for a screening tool. Output completeness, defined as the proportion of notes returning a parsable classification, was recorded for every model and inference configuration. All analyses were performed in Python (SciPy, statsmodels, pandas). Statistical significance was set at p < 0.05 (two-sided).

## Results

### Study Population

Of 3,023 triage notes analysed, 140 (4.6%) had a Code Stroke activation during the study period and 17 patients received a final stroke diagnosis without code activation. The neurologist reference standard identified 136 (4.5%) presentations as stroke-positive. Patients with a stroke code were older (median age 69 years, IQR 56–81) than those without (median 48 years, IQR 32–69), and 56.4% were male. Among the 140 code activations, 83 (59.2%) had a final diagnosis of stroke (74 ischaemic, 9 haemorrhagic), while 57 (40.7%) were ultimately determined not to have had a stroke. The most common non-stroke diagnoses among code activations were migraine (17.1%), peripheral vertigo (5.0%), seizure (4.3%), and delirium (4.3%). Median time from symptom onset was 5.9 hours (IQR 2.3–12.0) for code activations and 27.8 hours (IQR 16.4–71.5) for stroke diagnoses that did not receive a code. Endovascular clot retrieval was performed in 16 patients and thrombolysis in 4 (Table 1).

**Table 1.** Baseline Characteristics by Stroke Code Activation.

| Variable | Stroke Code | Final Stroke diagnosis & No Stroke Code | No Stroke diagnosis & No Code Activation |
| --- | --- | --- | --- |
| N | 140 | 17 | 2866 |
| Age, median (IQR) | 69 (56–81) | 75.0 (61.5–82.0) | 48.0 (32.0–69.0) |
| Male, n (%) | 79 (56.4%) | 9 (52.9%) |  |
| Neurologist label positive, n (%) | 105 (75.0%) | 3 (17.6%) | 28 (1.0%) |
| NIHSS, median (IQR) | 2.0 (0.0–7.0) | 2.0 (1.0–4.5) |  |
| Time from onset (hours), median (IQR) | 5.9 (2.3–12.0) | 27.8 (16.4–71.5) |  |
| Ischaemic | 74 (52.9%) | 17 (100.0%) |  |
| Haemorrhagic | 9 (6.4%) | 0 (0.0%) |  |
| No stroke | 57 (40.7%) | 0 (0.0%) |  |

| Final Diagnosis |  |  |  |
| --- | --- | --- | --- |
| Stroke | 83 (59.2%) | 17 (100.0%) |  |
| Migraine | 24 (17.1%) |  |  |
| Seizure | 6 (4.3%) |  |  |
| Peripheral Vertigo | 7 (5.0%) |  |  |
| Bell's Palsy | 2 (1.4%) |  |  |
| Delirium | 6 (4.3%) |  |  |
| Other | 12 (8.6%) |  |  |
| Occlusion Location |  |  |  |
| ICA-EC | 4 (2.9%) | 0 (0.0%) |  |
| ICA-IC | 2 (1.4%) | 0 (0.0%) |  |
| MCA-M1 | 12 (8.6%) | 0 (0.0%) |  |
| MCA-M2 | 5 (3.6%) | 1 (5.9%) |  |
| MCA-M3 | 1 (0.7%) | 0 (0.0%) |  |
| ACA | 1 (0.7%) | 0 (0.0%) |  |
| PCA | 0 (0.0%) | 0 (0.0%) |  |
| BA | 0 (0.0%) | 0 (0.0%) |  |
| VA | 0 (0.0%) | 0 (0.0%) |  |
| No occlusion | 115 (82.1%) | 16 (94.1%) |  |
| Intervention |  |  |  |
| ECR | 16 (11.4%) | 0 (0.0%) | 0 (0.0%) |
| tPA | 4 (2.9%) | 0 (0.0%) | 0 (0.0%) |

Values are median (IQR) or n (%).

### Reference Standard Reliability

In the blinded inter-rater sub-study, a second neurologist independently re-labelled 200 presentations (100 originally labelled stroke-positive and 100 stroke-negative). The two reviewers agreed on 190 of 200 presentations, giving a Cohen’s kappa of 0.900 (95% CI 0.838–0.959), corresponding to almost perfect agreement. Of the 10 disagreements, 8 were presentations labelled stroke-positive by the primary reviewer and stroke-negative by the blinded reviewer, and 2 were the reverse. This directional imbalance was not statistically significant (McNemar exact p = 0.11; bias index 0.03).

### Primary Model Comparison

Diagnostic performance of the six models using the prompt-chained pipeline is summarised in Figure 2. Against the neurologist reference standard, Qwen 2.5 14B achieved the highest sensitivity at 89.0% (95% CI 82.6–93.2%) with a specificity of 99.3% (98.9–99.6%), a positive predictive value of 85.8% (79.1–90.6%), and a negative predictive value of 99.5% (99.1–99.7%). Balanced accuracy ranged from 0.884 (Llama 3.1 8B) to 0.941 (Qwen 2.5 14B). Gwet’s AC1 indicated high agreement with the neurologist label across all models (range 0.938–0.987). Model performance was similar vs actual ED code activation summarised in supplementary Figure 1. Two-by-two counts and full performance estimates for each model are shown in Supplemental Table 1.

**Figure 2:**
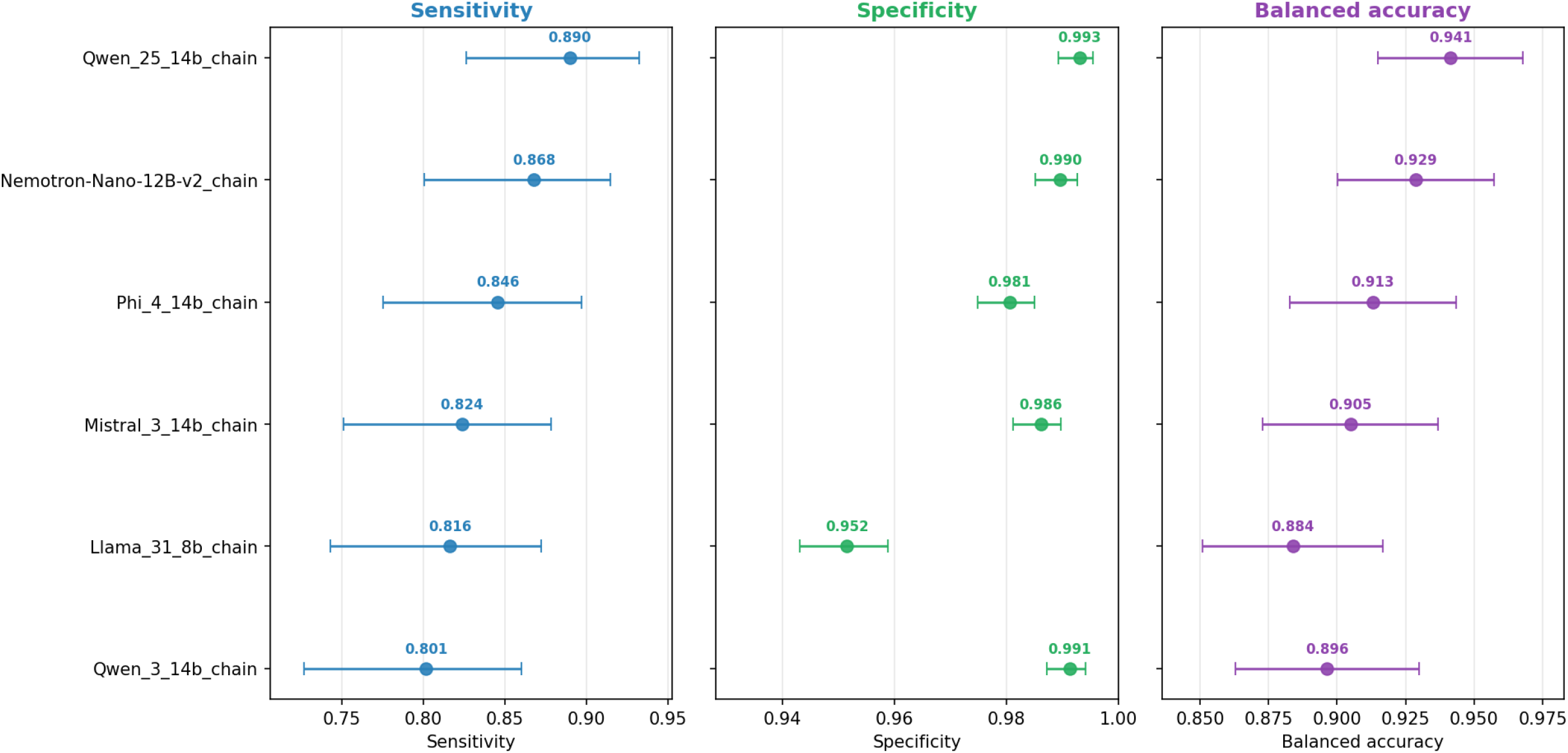
Per-model (chain design) diagnostic performance vs Neurologist label (Wilson 95% CI for sens/spec, normal-approx 95% CI for balanced acc.)

### Pairwise Model Comparisons

McNemar’s test with Holm correction identified several statistically significant differences in overall accuracy (Figure 3). Qwen 2.5 14B was significantly more accurate than Llama 3.1 8B (p < 0.001), Mistral 3 14B (p < 0.001), and Phi-4 14B (p < 0.001), but did not differ significantly from Nemotron-Nano-12B-v2 (p = 0.42) or Qwen 3 14B (p = 0.19). No pairwise comparisons reached significance for sensitivity alone after Holm correction.

**Figure 3:**
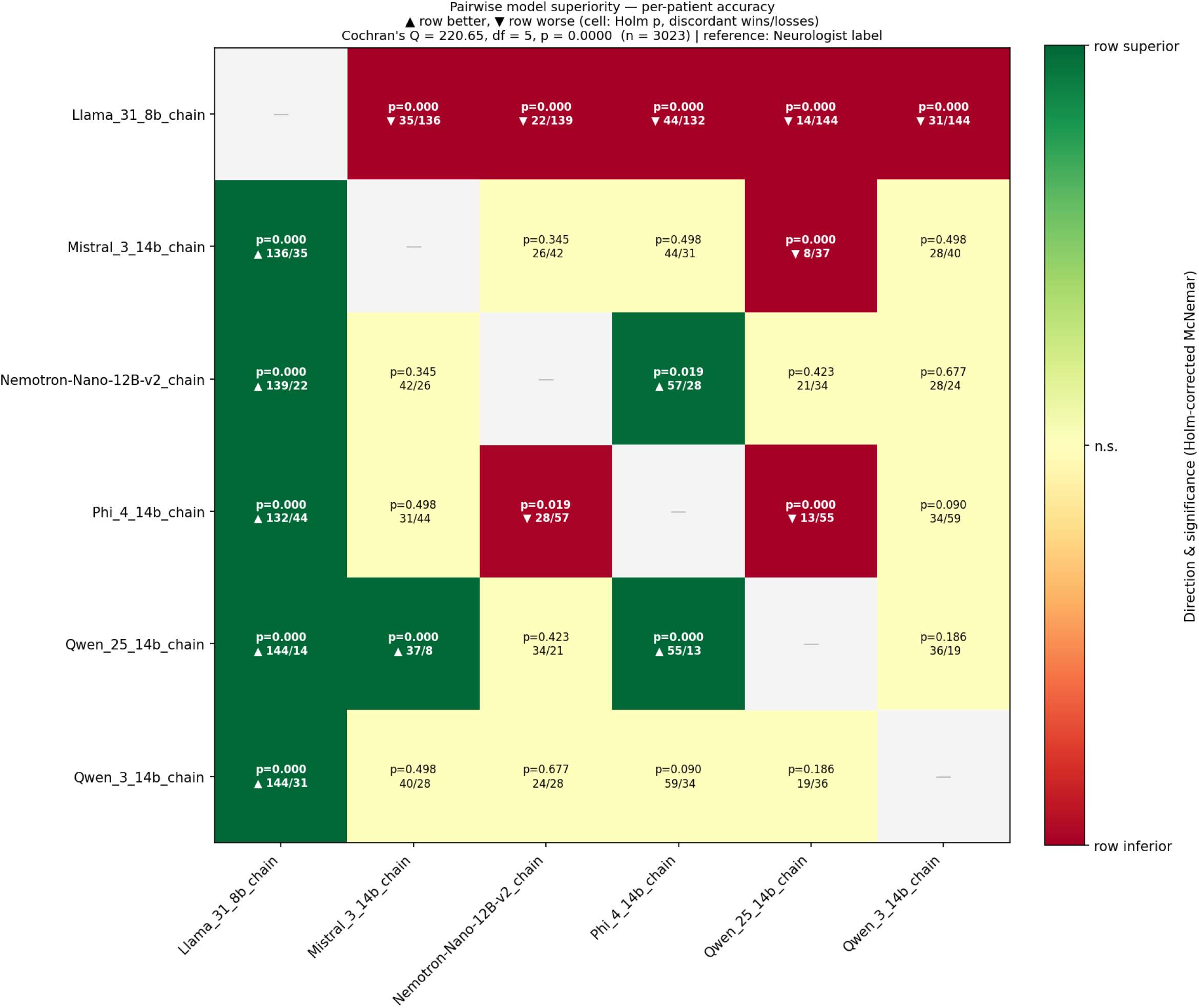
Pairwise Model Comparisons - McNemar’s test with Holm correction.

### Secondary Comparison: Prompt Chaining versus Single-Pass Inference

The prompt chaining architecture significantly improved overall accuracy for two of the three models tested. For Qwen 2.5 14B, the chain configuration achieved a sensitivity of 89.0% and specificity of 99.3%, compared with 77.2% and 98.5% respectively for the single-pass configuration (McNemar p < 0.001). For Llama 3.1 8B, the single-pass configuration achieved a higher sensitivity (98.5% versus 81.6%) but at the cost of a dramatically lower specificity (58.1% versus 95.2%), producing 1,210 false positives compared with 140 for the chain (p < 0.001). Two-by-two counts and full diagnostic performance estimates for each model and inference configuration are shown in Supplemental Table 2.

Qwen 3 14B showed no significant difference between chain and single-pass configurations in overall accuracy (p = 1.00). For this model, single-pass inference produced slightly higher sensitivity (83.8% versus 80.1%) with marginally lower specificity (98.8% versus 99.1%). Enabling thinking mode on the single-pass configuration for Qwen 3 14B did not significantly alter performance compared with either the chain (p = 1.00) or single-pass without thinking (p = 1.00) configurations (Figure 4).

**Figure 4:**
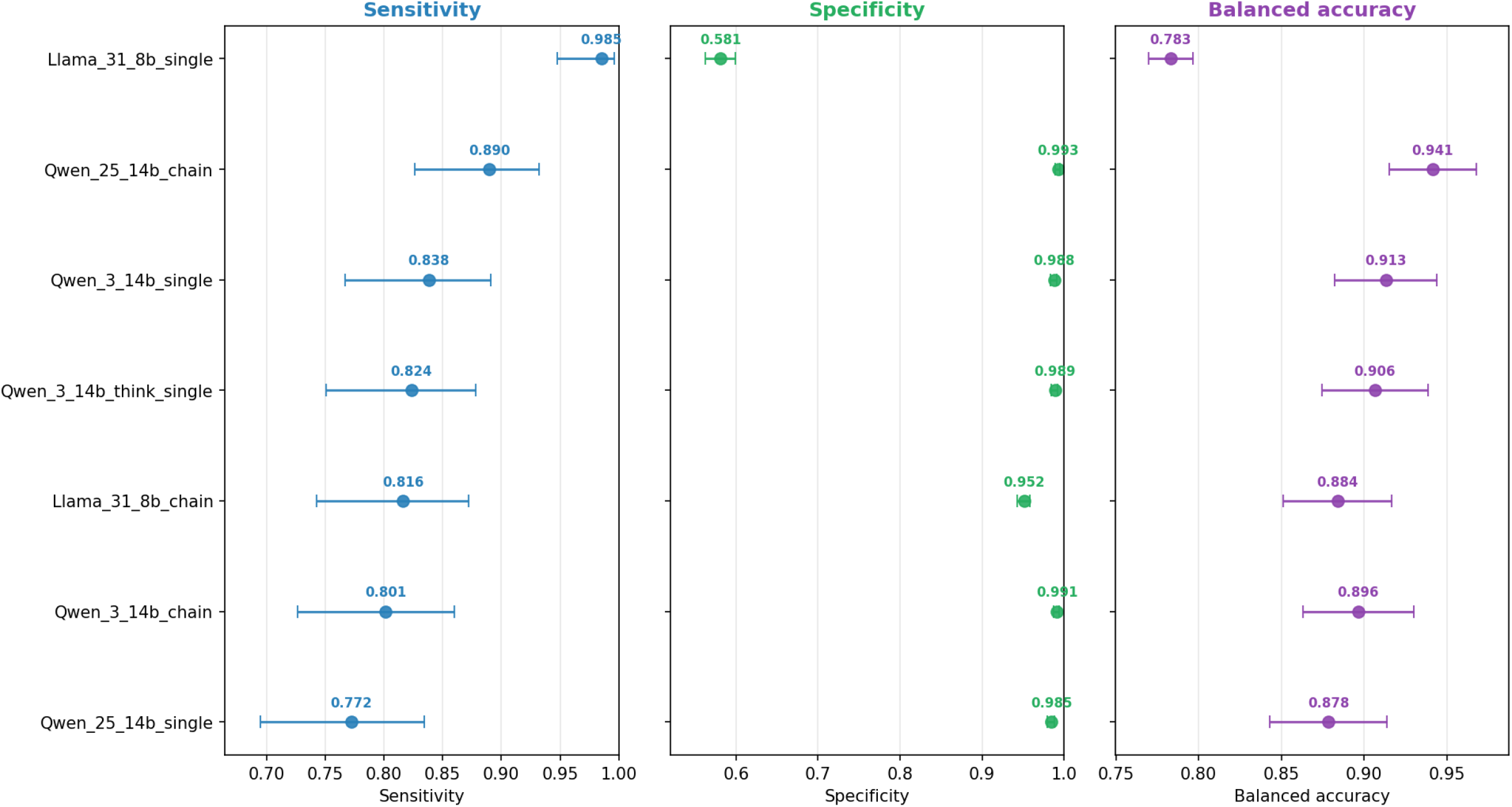
Per-model (single vs chain design) diagnostic performance vs Neurologist label (Wilson 95% CI for sens/spec, normal-approx 95% CI for balanced acc.)

### Output Completeness

Output completeness differed markedly between the two inference architectures. Across all six models, the multi-pass chain returned a complete, parsable classification for every note (3,023/3,023; 100%), with no missing or malformed output. Single-pass inference returned no usable classification for 90 of 3,023 notes with Llama 3.1 8B (3.0%), 37 with Qwen 2.5 14B (1.2%) and 2 with Qwen 3 14B (0.1%); enabling thinking mode on the Qwen 3 14B single-pass configuration produced 6 missing outputs (0.2%). All missing predictions were imputed as model-negative, so single-pass sensitivity estimates are lower bounds and single-pass specificity estimates upper bounds.

## Discussion

We demonstrated that a prompt-chained pipeline using lightweight, locally deployed language models identified presentations meeting institutional Code Stroke criteria with high diagnostic accuracy against a blinded neurologist reference standard. The best-performing configuration (Qwen 2.5 14B with the multi-pass chain) achieved a sensitivity of 89.0% and specificity of 99.3% against a neurologist reference standard. These results were achieved using general-purpose pre-trained models of 8–14 billion parameters running on a single consumer-grade GPU, without fine-tuning, cloud connectivity, or exposure of patient data to any external system.

The secondary comparison demonstrates that prompt architecture materially changes the clinical safety profile of an LLM screening tool. Collapsing all clinical reasoning into a single inference call degraded performance in model-specific and clinically consequential ways. For Llama 3.1 8B, single-pass inference inflated sensitivity to 98.5% while collapsing specificity to 58.1%, generating over 1,200 false positives. Enabling the internal reasoning (“thinking”) mode of Qwen 3 14B did not significantly improve performance over deterministic decoding, while sacrificing run-to-run reproducibility (important for clinical audit and regulatory evaluation). Using an early exit architecture ensured that the chain pipeline did not result in excessive resource utilisation and pipeline delay. Chain prompting pipelines have been previously shown to reduce hallucination and this was indeed evident in our real-world application.

The deployment architecture addresses two key barriers to clinical LLM adoption. First, the entire pipeline operates on device without internet connectivity, eliminating external transmission of patient data and simplifying data sovereignty compliance. Second, because the models are general-purpose and receive no training on patient data, no patient information can be internalised in model weights. This avoids both privacy risk and the regulatory complexity of locally trained models. The pipeline is also readily transferable to other hospital environments. Translation prompts can be adapted to incorporate local clinical abbreviations and social conventions, allowing institutions to tune performance without model retraining. New centres would need only to deploy the audit and optimisation pipeline to evaluate how prompt modifications affect performance. This is far simpler than undertaking model training from scratch. Combined with a hardware requirement of just a single consumer GPU, this approach is accessible to individual departments without the need for enterprise infrastructure.

Several limitations should be acknowledged. This was a single-centre retrospective study over a one-month period; performance may differ across institutions with different documentation cultures, triage practices, and patient populations, and prospective validation is required before clinical deployment. Reliability of the neurologist label was supported by a blinded sub-study in which a second neurologist showed almost perfect agreement with the primary reviewer (κ = 0.900); however, agreement was assessed on a stratified sub-sample rather than the full cohort, on the composite binary label rather than its individual components, and between two reviewers applying the same institutional criteria, so systematic error common to both reviewers cannot be excluded. Triage notes were in English with Australian clinical shorthand, and the translation step was tailored accordingly; generalisation to other linguistic contexts is untested. Inference speed was not optimised or formally evaluated, although the sequential architecture and modest hardware suggest per-note processing times compatible with triage workflows. Finally, the stroke prevalence in this cohort (4.5%) reflects an undifferentiated ED population; positive predictive value will vary with local prevalence.

The next step is prospective evaluation whilst embedded in the live triage workflow, measuring processing latency, alert burden, and the effect of automated flagging on staff behaviours, door-to-needle and door-to-groin times. Multi-site validation across varied documentation styles and demographic case-mixes would establish generalisability. The pipeline’s component-level structure lends itself to targeted refinement as new failure modes are identified. Beyond technical performance, sustainable deployment will require streamlined support infrastructure (robust auditing, maintenance workflows, and clear governance) before these tools can be responsibly embedded in clinical practice.

## Conclusion

A prompt-chained pipeline of lightweight, locally deployed, general-purpose language models identified Code Stroke-eligible presentations from free-text ED triage notes with high sensitivity and specificity. It operated entirely on device with no patient data leaving the hospital. Prompt architecture proved as consequential as model selection: chained, deterministic inference substantially outperformed single pass approaches whose failure modes were clinically unacceptable. These findings support prospective evaluation of locally deployed LLM screening as a privacy-preserving safety net at ED triage.

## Data Availability

All data produced in the present study are available upon reasonable request to the authors

## References

[1] V. L. Feigin et al., “World stroke organization: global stroke fact sheet 2025,” International Journal of Stroke, vol. 20, no. 2, pp. 132–144, 2025.

[2] M. Hilditch, C. Brand, S. Devlin, A. Boyd, and E. Venema, “BE-FAST vs FAST in prehospital stroke recognition: a systematic review,” British Journal of Community Nursing, vol. 30, no. 9, pp. 439–447, 2025.

[3] M. J. Mulder et al., “Time to endovascular treatment and outcome in acute ischemic stroke: MR CLEAN registry results,” Circulation, vol. 138, no. 3, pp. 232–240, 2018.

[4] L. Ma et al., “The third Intensive Care Bundle with Blood Pressure Reduction in Acute Cerebral Haemorrhage Trial (INTERACT3): an international, stepped wedge cluster randomised controlled trial,” The Lancet, vol. 402, no. 10395, pp. 27–40, 2023.

[5] D. E. Newman-Toker, E. Moy, E. Valente, R. Coffey, and A. L. Hines, “Missed diagnosis of stroke in the emergency department: a cross-sectional analysis of a large population-based sample,” Diagnosis, vol. 1, no. 2, pp. 155–166, 2014.

[6] M. F. Gude et al., “Factors associated with missed stroke diagnosis in prehospital triage: a single-center cohort study,” BMC neurology, 2026.

[7] B. Yan et al., “On protecting the data privacy of Large Language Models (LLMs) and LLM agents: A literature review,” High-Confidence Computing, vol. 5, no. 2, p. 100300, 2025.

[8] C. Zhang, H. Wang, and H. Meng, “Hallucination detection and evaluation of large language model,” arXiv preprint arXiv:2512.22416, 2025.

[9] L. Barkley and B. van der Merwe, “Investigating the role of prompting and external tools in hallucination rates of large language models,” arXiv preprint arXiv:2410.19385, 2024.

[10] C. Wang, Y. Wu, and J. Zhong, “Mitigating Implicit Hallucinations in Large Language Models Based on Progressive Prompt Chains,” 2025.

[11] H. Zhou, “From Hallucination to Structure Snowballing: The Alignment Tax of Constrained Decoding in LLM Reflection,” arXiv preprint arXiv:2604.06066, 2026.

[12] M. Bujang and N. Baharim, “Guidelines of the minimum sample size requirements for Cohen’s Kappa. Epidemiol,” Biostatistics Public Health, vol. 14, pp. e12267–12261, 2017.

