## Supplemental Figure 1 for "Diagnostic Accuracy of a locally deployed Large Language Model Algorithm for Automated Code Stroke Pathway Identification in Emergency Department Triage Notes"

**Supplemental Figure 1: Per-model (chain design) diagnostic performance vs Actual ED code activation (Wilson 95% CI for sens/spec, normal-approx 95% CI for balanced acc.)**
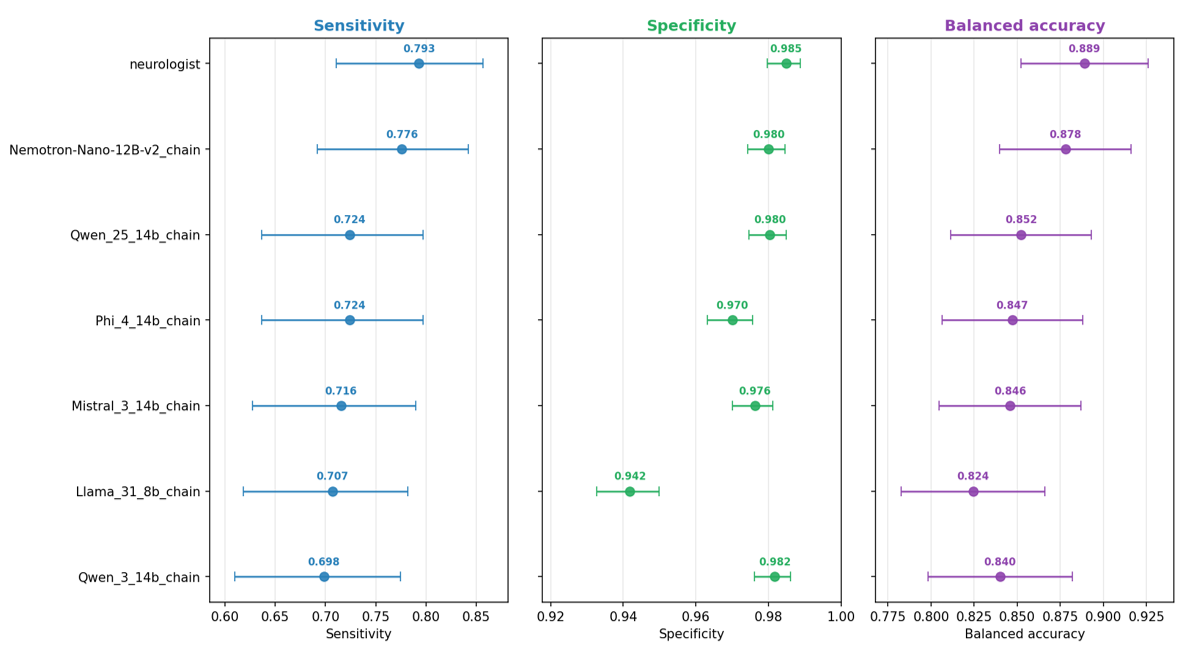
