## Supplemental Table 1 for "Diagnostic Accuracy of a locally deployed Large Language Model Algorithm for Automated Code Stroke Pathway Identification in Emergency Department Triage Notes"

**Table 1. Two-by-two counts and diagnostic performance of the multi-pass chain pipeline by model, against the neurologist reference standard**

| **Model** | **TP** | **FP** | **FN** | **TN** | **Sensitivity, % (95% CI)** | **Specificity, % (95% CI)** | **PPV, % (95% CI)** | **NPV, % (95% CI)** | **Balanced accuracy (95% CI)** | **Cohen's κ** | **Gwet's AC1** |
| --- | --- | --- | --- | --- | --- | --- | --- | --- | --- | --- | --- |
| Qwen 2.5 14B | 121 | 20 | 15 | 2867 | 89.0 (82.6–93.2) | 99.3 (98.9–99.6) | 85.8 (79.1–90.6) | 99.5 (99.1–99.7) | 0.941 (0.915–0.968) | 0.868 | 0.987 |
| Nemotron-Nano-12B-v2 | 118 | 30 | 18 | 2857 | 86.8 (80.0–91.5) | 99.0 (98.5–99.3) | 79.7 (72.5–85.4) | 99.4 (99.0–99.6) | 0.929 (0.900–0.957) | 0.823 | 0.983 |
| Phi-4 14B | 115 | 56 | 21 | 2831 | 84.6 (77.5–89.7) | 98.1 (97.5–98.5) | 67.3 (59.9–73.8) | 99.3 (98.9–99.5) | 0.913 (0.883–0.944) | 0.736 | 0.972 |
| Mistral 3 14B | 112 | 40 | 24 | 2847 | 82.4 (75.1–87.8) | 98.6 (98.1–99.0) | 73.7 (66.2–80.0) | 99.2 (98.8–99.4) | 0.905 (0.873–0.937) | 0.767 | 0.977 |
| Llama 3.1 8B | 111 | 140 | 25 | 2747 | 81.6 (74.3–87.2) | 95.2 (94.3–95.9) | 44.2 (38.2–50.4) | 99.1 (98.7–99.4) | 0.884 (0.851–0.917) | 0.547 | 0.938 |
| Qwen 3 14B | 109 | 25 | 27 | 2862 | 80.1 (72.7–86.0) | 99.1 (98.7–99.4) | 81.3 (73.9–87.0) | 99.1 (98.6–99.4) | 0.896 (0.863–0.930) | 0.798 | 0.981 |

*TP, true positive; FP, false positive; FN, false negative; TN, true negative; PPV, positive predictive value; NPV, negative predictive value. All estimates are against the neurologist reference standard (136 positive of 3,023 presentations). Sensitivity, specificity, PPV and NPV are shown with Wilson score 95% confidence intervals; balanced accuracy with a normal-approximation 95% confidence interval. Models are ordered by sensitivity. The chain returned a parsable classification for every note in every model (3,023/3,023).*
