## Supplemental Table 2 for "Diagnostic Accuracy of a locally deployed Large Language Model Algorithm for Automated Code Stroke Pathway Identification in Emergency Department Triage Notes"

**Table 2. Two-by-two counts and diagnostic performance by model and inference configuration, against the neurologist reference standard**

| **Model and configuration** | **TP** | **FP** | **FN** | **TN** | **Sensitivity, % (95% CI)** | **Specificity, % (95% CI)** | **PPV, % (95% CI)** | **NPV, % (95% CI)** | **Balanced accuracy** | **Cohen's κ** | **Gwet's AC1** |
| --- | --- | --- | --- | --- | --- | --- | --- | --- | --- | --- | --- |
| Llama 3.1 8B — chain | 111 | 140 | 25 | 2747 | 81.6 (74.3–87.2) | 95.2 (94.3–95.9) | 44.2 (38.2–50.4) | 99.1 (98.7–99.4) | 0.884 | 0.547 | 0.938 |
| Llama 3.1 8B — single-pass | 134 | 1210 | 2 | 1677 | 98.5 (94.8–99.6) | 58.1 (56.3–59.9) | 10.0 (8.5–11.7) | 99.9 (99.6–100.0) | 0.783 | 0.108 | 0.364 |
| Qwen 2.5 14B — chain | 121 | 20 | 15 | 2867 | 89.0 (82.6–93.2) | 99.3 (98.9–99.6) | 85.8 (79.1–90.6) | 99.5 (99.1–99.7) | 0.941 | 0.868 | 0.987 |
| Qwen 2.5 14B — single-pass | 105 | 44 | 31 | 2843 | 77.2 (69.5–83.5) | 98.5 (98.0–98.9) | 70.5 (62.7–77.2) | 98.9 (98.5–99.2) | 0.878 | 0.724 | 0.973 |
| Qwen 3 14B — chain | 109 | 25 | 27 | 2862 | 80.1 (72.7–86.0) | 99.1 (98.7–99.4) | 81.3 (73.9–87.0) | 99.1 (98.6–99.4) | 0.896 | 0.798 | 0.981 |
| Qwen 3 14B — single-pass | 114 | 35 | 22 | 2852 | 83.8 (76.7–89.1) | 98.8 (98.3–99.1) | 76.5 (69.1–82.6) | 99.2 (98.8–99.5) | 0.913 | 0.790 | 0.979 |
| Qwen 3 14B — single-pass, thinking | 112 | 31 | 24 | 2856 | 82.4 (75.1–87.8) | 98.9 (98.5–99.2) | 78.3 (70.9–84.3) | 99.2 (98.8–99.4) | 0.906 | 0.793 | 0.980 |

*TP, true positive; FP, false positive; FN, false negative; TN, true negative; PPV, positive predictive value; NPV, negative predictive value. All estimates are against the neurologist reference standard (136 positive of 3,023 presentations). Sensitivity, specificity, PPV and NPV are shown with Wilson score 95% confidence intervals. Missing or unparsable model outputs were imputed as model-negative.*
